## Supplemental Figures and Tables for "Boosting of Cross-Reactive Antibodies to Endemic Coronaviruses by SARS-CoV-2 Infection but not Vaccination with Stabilized Spike"

**Supplementary Materials**

| **Supplementary Figures** | |
|---|---|
| Supplementary Figure 1 | Structural differences between the S1 domain of SARS-CoV-2 and endemic strains. |
| Supplementary Figure 2 | IgA and IgM responses in the DHMC cohort. |
| Supplementary Figure 3 | Antibody responses in the JHMI cohort. |
| Supplementary Figure 4 | Elevated IgG but not IgM responses to endemic CoV in convalescent cohorts. |
| Supplementary Figure 5 | Elevated responses to endemic CoV in the pre- and post-infection cohort. |
| Supplementary Figure 6 | Correlative relationships between SARS-CoV-2- and OC43-specific antibody responses in the JHMI cohort by isotype. |
| Supplementary Figure 7 | Cross-reactivity of SARS-CoV-2 S-2P specific IgG. |
| Supplementary Figure 8 | Cross-reactivity of SARS-CoV-2 RBD specific IgG. |
| Supplementary Figure 9 | Cross-reactivity of SARS-CoV-2 S2 specific IgG. |
| Supplementary Figure 10 | Cross-reactivity of OC43 S specific IgG. |
| Supplementary Figure 11 | Cross-reactivity of SARS-CoV-2 S-2P specific IgA. |
| Supplementary Figure 12 | Cross-reactivity of SARS-CoV-2 RBD specific IgA. |
| Supplementary Figure 13 | Cross-reactivity of SARS-CoV-2 S2 specific IgA. |
| Supplementary Figure 14 | Cross-reactivity of OC43 S specific IgA. |
| Supplementary Figure 15 | Cross-reactivity of SARS-CoV-2 S-2P specific IgM. |
| Supplementary Figure 16 | Cross-reactivity of SARS-CoV-2 RBD specific IgM. |
| Supplementary Figure 17 | Cross-reactivity of SARS-CoV-2 S2 specific IgM. |
| Supplementary Figure 18 | Cross-reactivity of OC43 S specific IgM. |
| Supplementary Figure 19 | IgA and IgM responses among naïve subjects, infected pregnant women, vaccinated adults, and vaccinated pregnant women. |
| **Supplementary Tables** | |
| Supplementary Table 1 | Coronavirus structures. |
| Supplementary Table 2 | Fc detection and antigen reagents. |

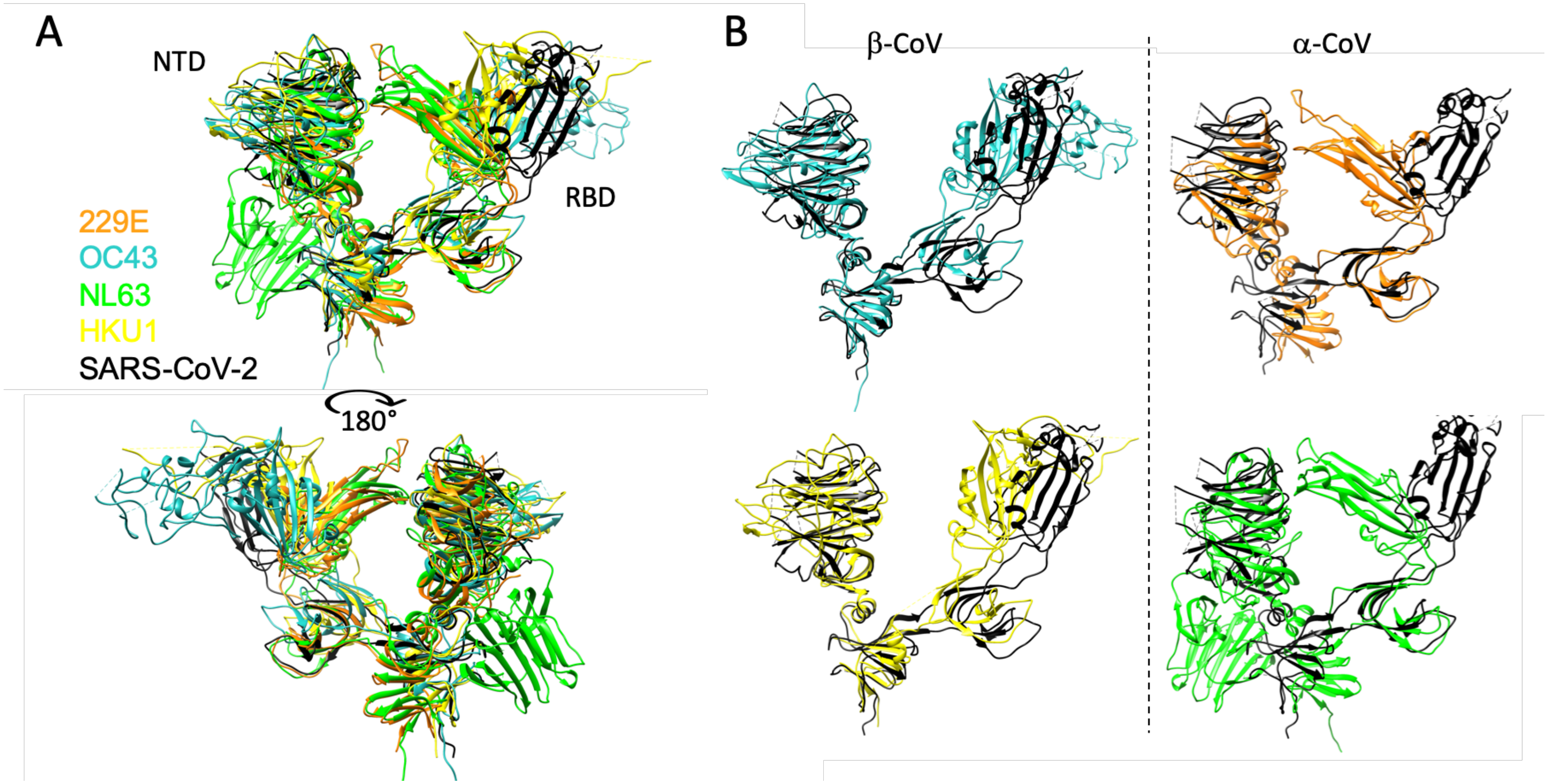


**Supplemental Figure 1. Structural differences between the S1 domain of SARS-CoV-2 and endemic strains. A**. Structural models of the NTD and RBD as ribbons, colored by strain: SARS-CoV-2 (black), 229E (orange), OC43 (blue), NL63 (green), and HKU1 (yellow). Structural alignments were restricted to the residues of the S1 domain. Right- alignments between SARS-CoV2 and the endemic S1 domains shown individually rather than overlayed, with β-CoV at left, and α-CoV at right.

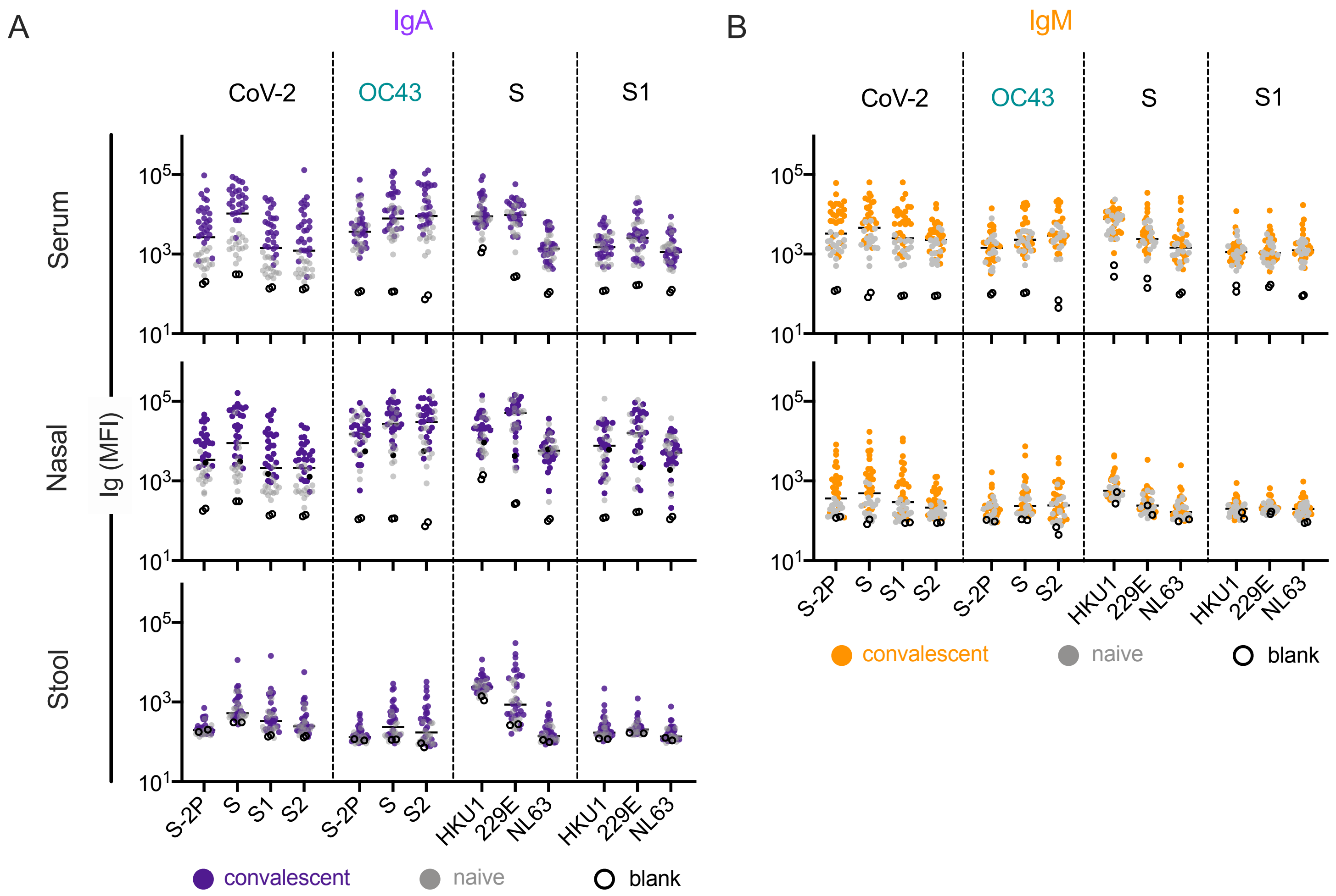


**Supplemental Figure 2. IgA and IgM responses in the DHMC cohort**. **A-B**. IgA (**A**) and IgM (**B**) responses in serum (top), nasal wash (middle) and stool (bottom) across antigens from CoV-2, OC43, and other endemic CoV S, and S1 proteins. Samples from naïve subjects are indicated in gray, SARS-CoV-2 convalescents at one month post infection in color, and buffer blanks in hollow circles.

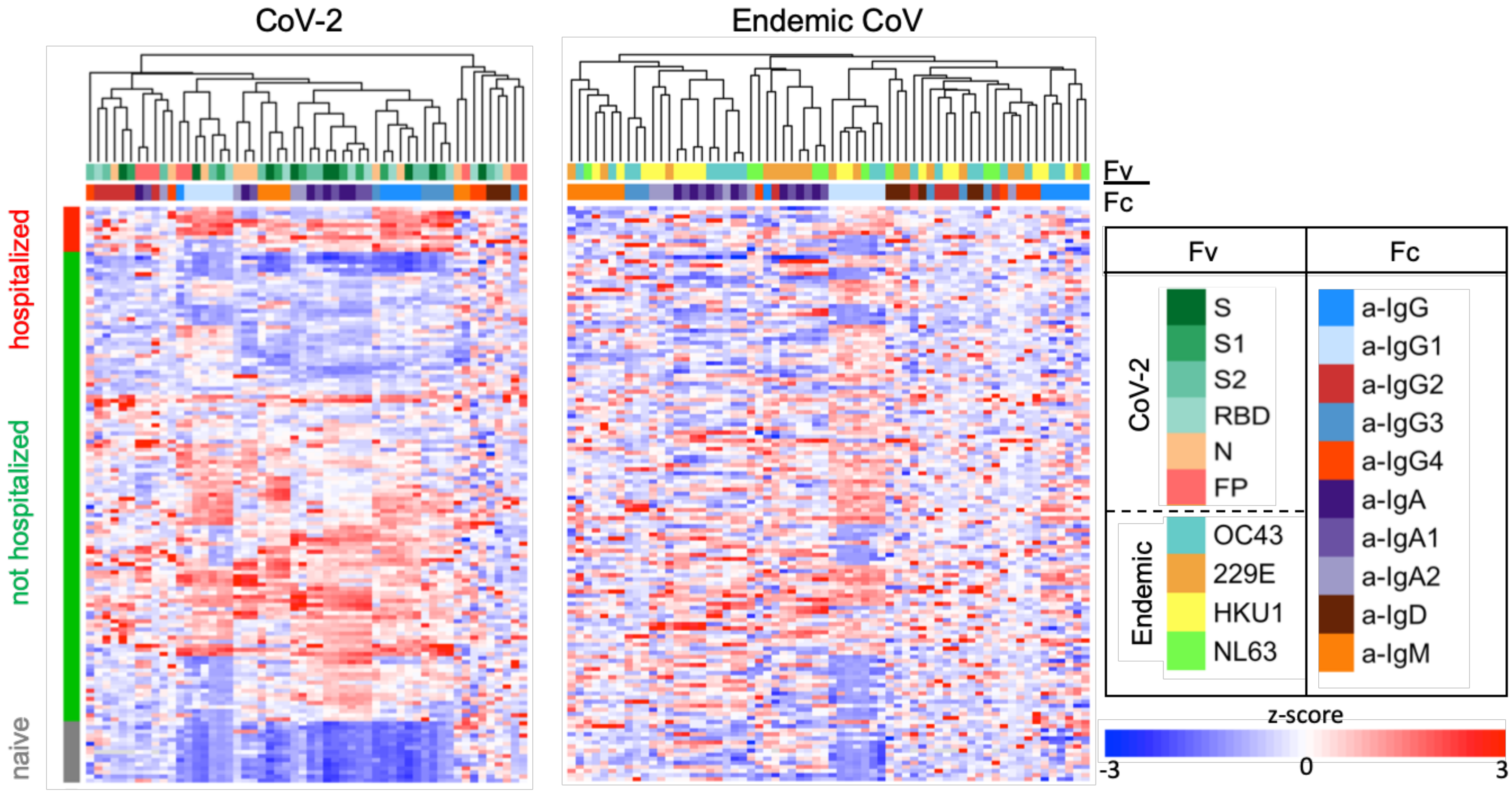


**Supplemental Figure 3. Antibody responses in the JHMI cohort**. Heatmap of filtered and hierarchically clustered features and within subject groups according to infection and hospitalization status. Ab responses to SARS CoV-2 features are shown on the left and those specific to endemic CoV on the right. Responses were scaled and centered within features and the scale was truncated at +/- 3 SD. Antigen specificity (Fv) and Fc characteristics (Fc) are indicated in the color bars.

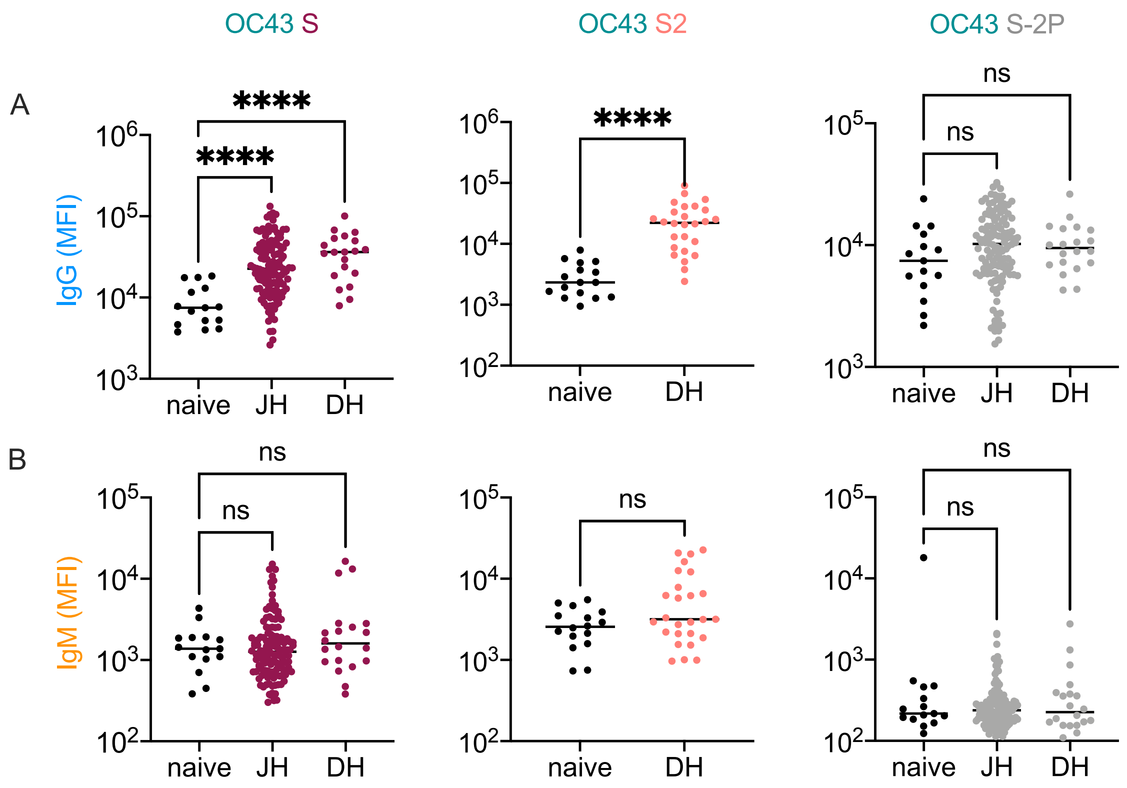


**Supplemental Figure 4. Elevated IgG but not IgM responses to endemic CoV in convalescent cohorts**. **A-B**. Comparison between IgG (**A**) and IgM (**B**) levels in naïve, DHMC (DH), and JHMI (JH) cohort samples to OC43 S, OC43 S2, and OC43 S-2P. Significant differences were defined by ANOVA with Dunnett's correction (****$p<0.0001$).

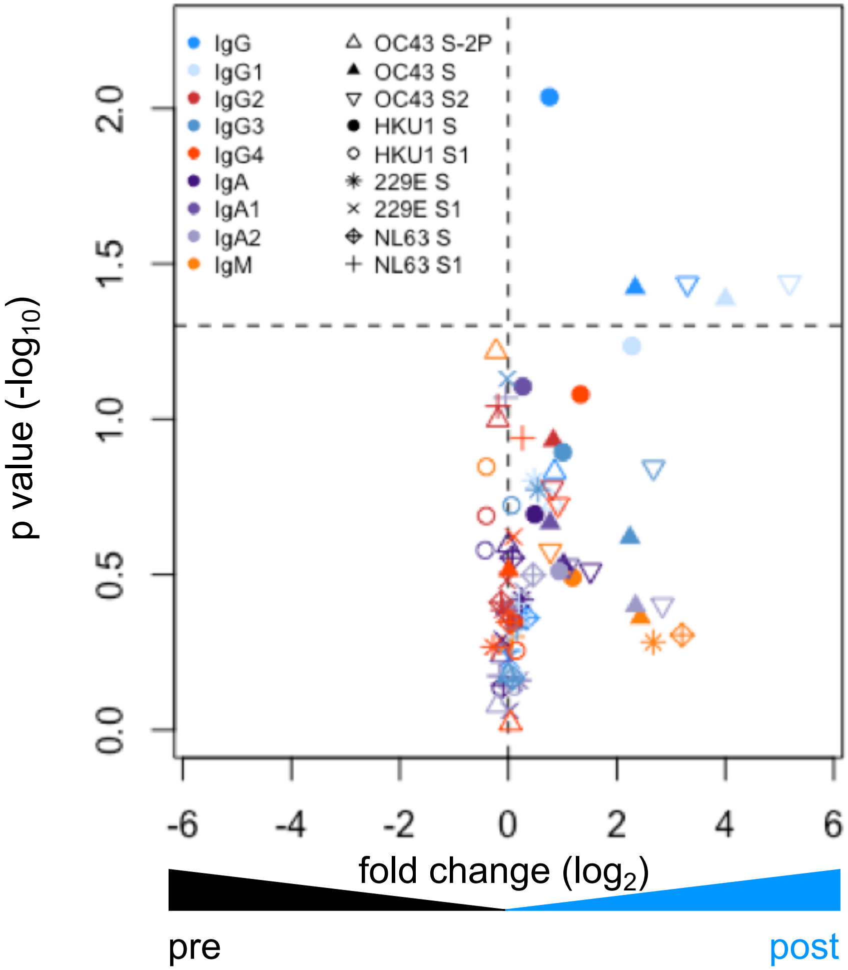


**Supplemental Figure 5. Elevated responses to endemic CoV in the pre- and post-infection cohort**. Volcano plot of fold change and significance (paired t test) of differences between antibody responses observed in convalescent subjects in the pre- and post-infection cohort. Dotted horizontal line indicates unadjusted p = 0.05. Each symbol represents an antibody response feature, with Fc domain characteristics represented by color and Fv antigen-specificity indicated by shape.

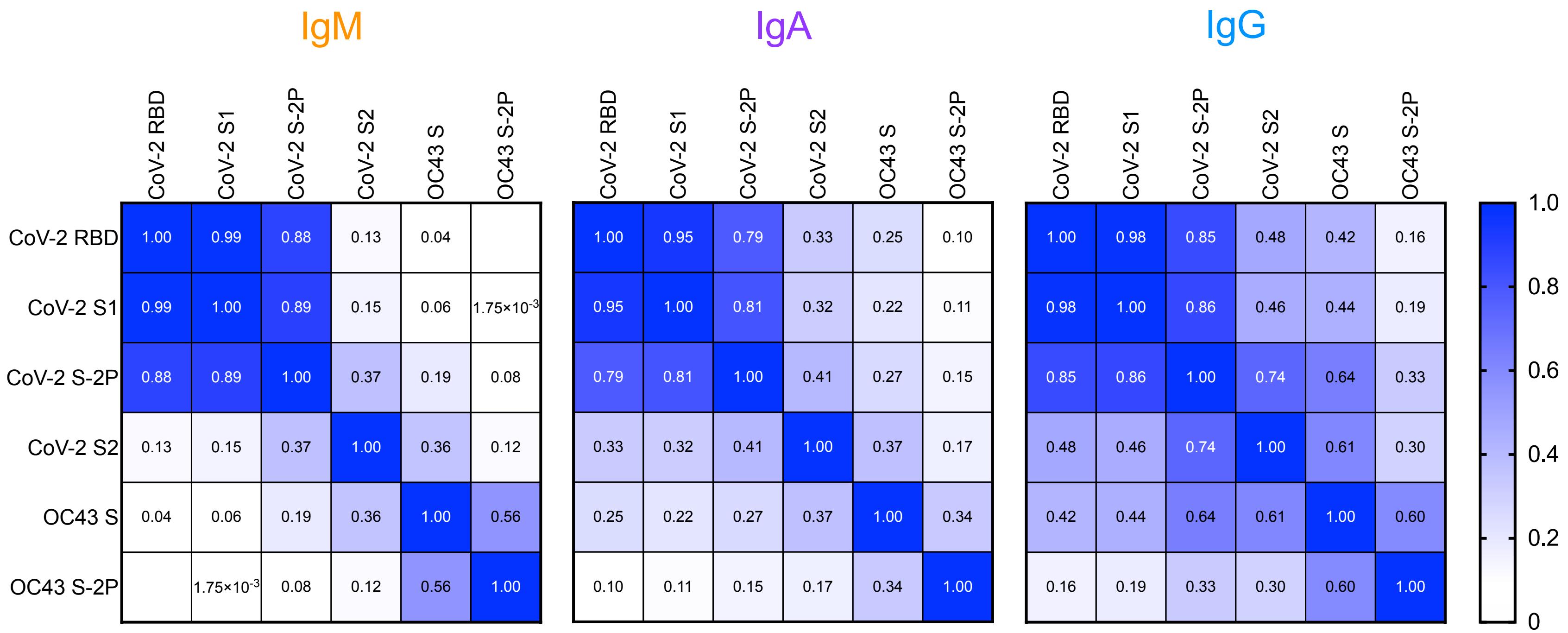


**Supplemental Figure 6. Correlative relationships between SARS-CoV-2- and OC43-specific antibody responses in the JHMI cohort by isotype**. Heatmap of Pearson correlation coefficients observed among SARS-CoV-2- and OC43-specific IgM (left), IgA (center), and IgG (right) isotypes.

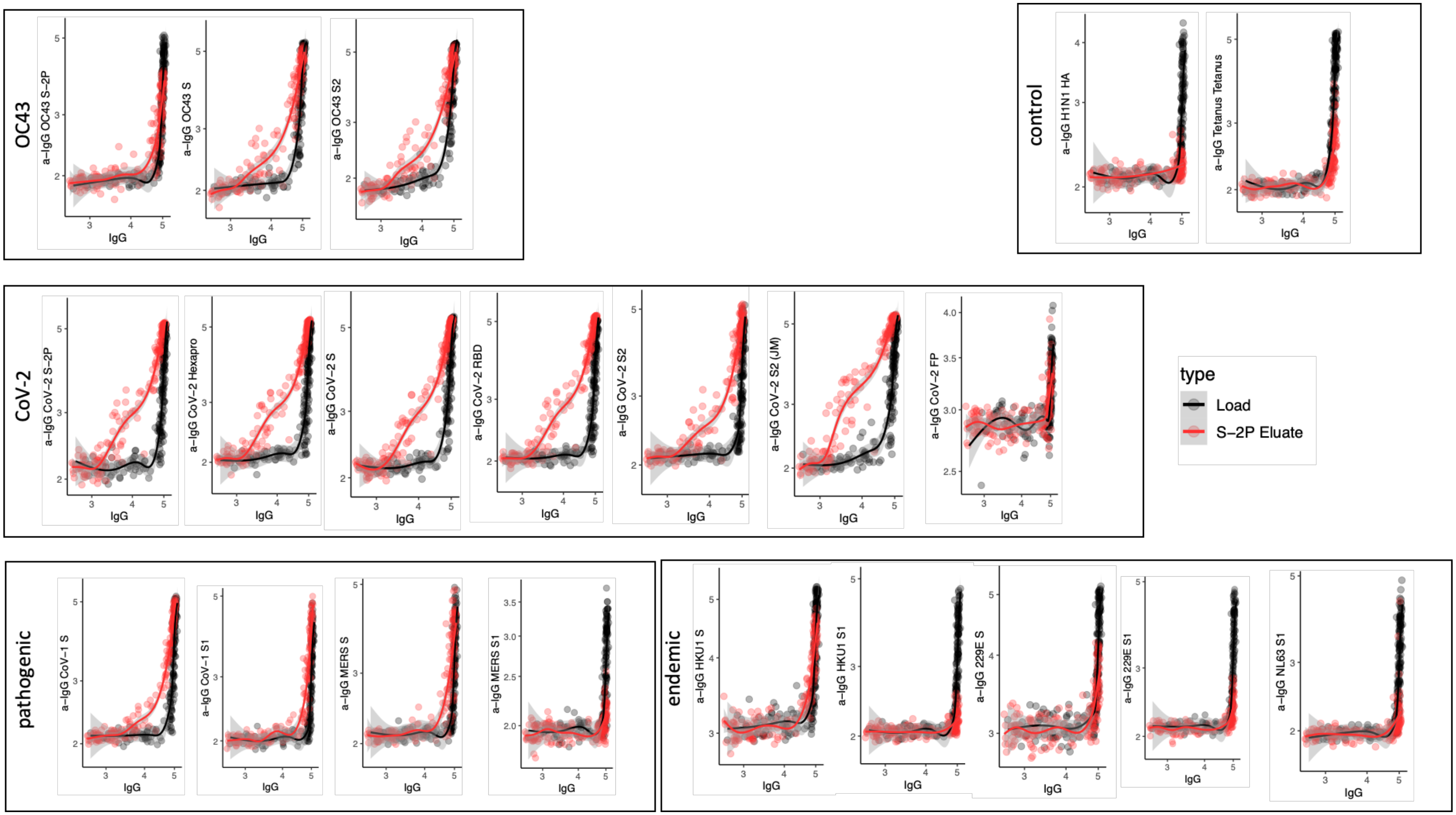


**Supplemental Figure 7: Cross-reactivity of SARS-CoV-2 S-2P specific IgG.** Antigen binding profiles of IgG in unfractionated serum (load, black) and affinity-purified CoV-2 S-2P- (eluate, red) fractions from 30 SARS-CoV-2 convalescent subjects across OC43, control, CoV-2, pathogenic CoV, and other endemic CoV. Y-axis depicts binding signal for indicated antigen specificity, x-axis depicts signal from total IgG quantitation.

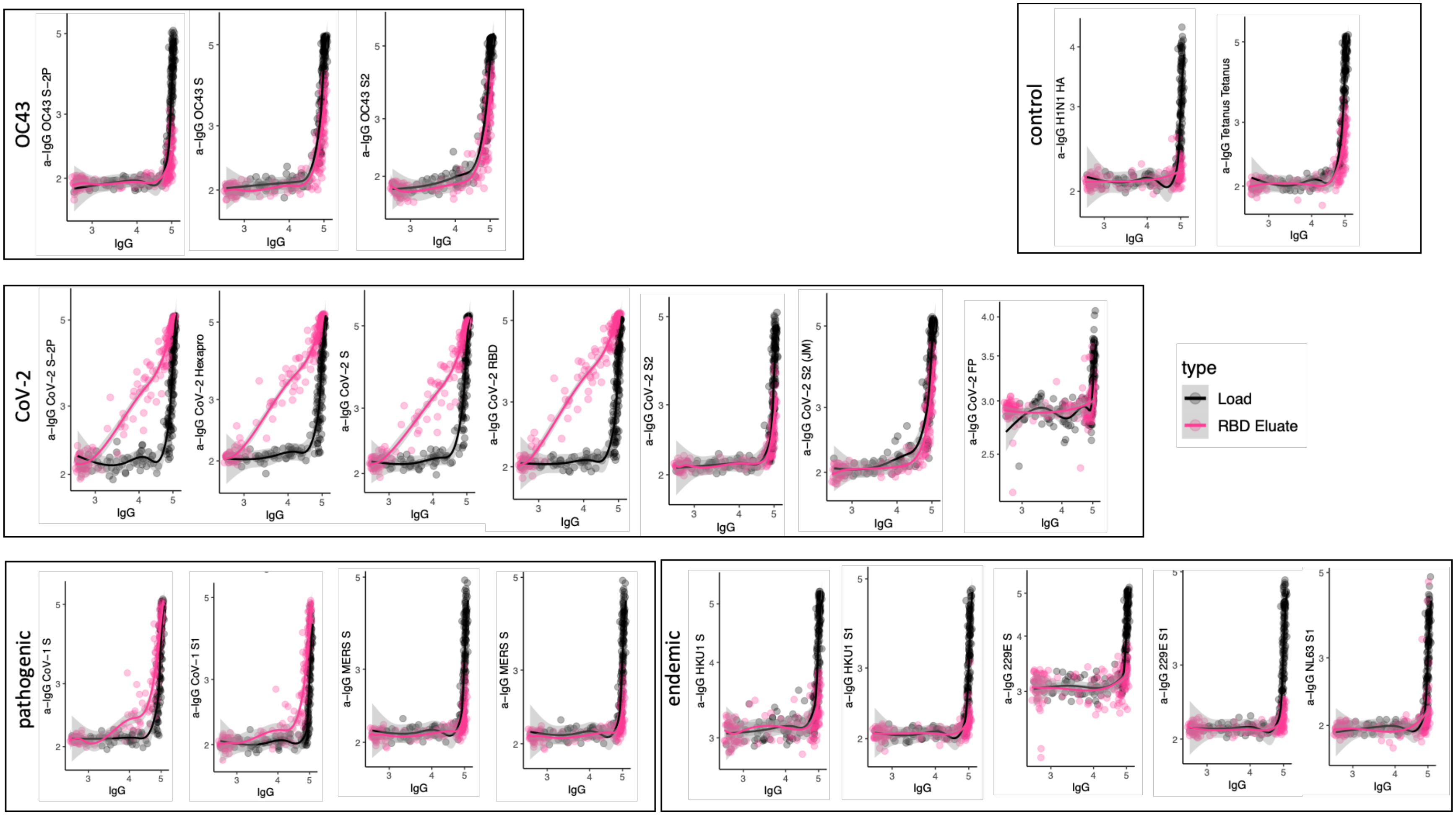


**Supplemental Figure 8: Cross-reactivity of SARS-CoV-2 RBD specific IgG.** Antigen binding profiles of IgG in unfractionated serum (load, black) and affinity-purified CoV-2 RBD- (eluate, pink) fractions from 30 SARS-CoV-2 convalescent subjects across OC43, control, CoV-2, pathogenic CoV, and other endemic CoV. Y-axis depicts binding signal for indicated antigen specificity, x-axis depicts signal from total IgG quantitation.

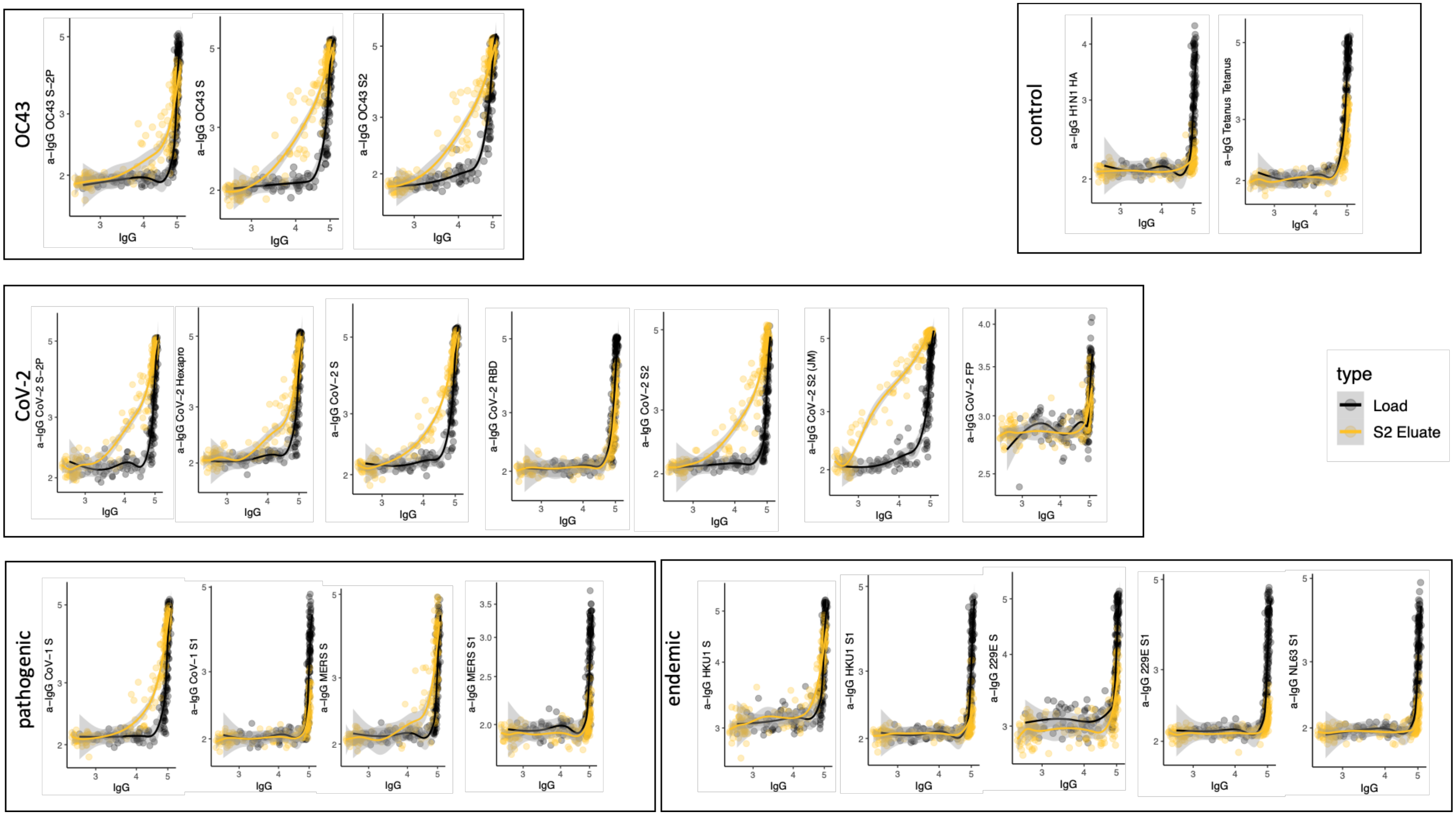


**Supplemental Figure 9: Cross-reactivity of SARS-CoV-2 S2 specific IgG.** Antigen binding profiles of IgG in unfractionated serum (load, black) and affinity-purified CoV-2 S2- (eluate, yellow) fractions from 30 SARS-CoV-2 convalescent subjects across OC43, control, CoV-2, pathogenic CoV, and other endemic CoV. Y-axis depicts binding signal for indicated antigen specificity, x-axis depicts signal from total IgG quantitation.

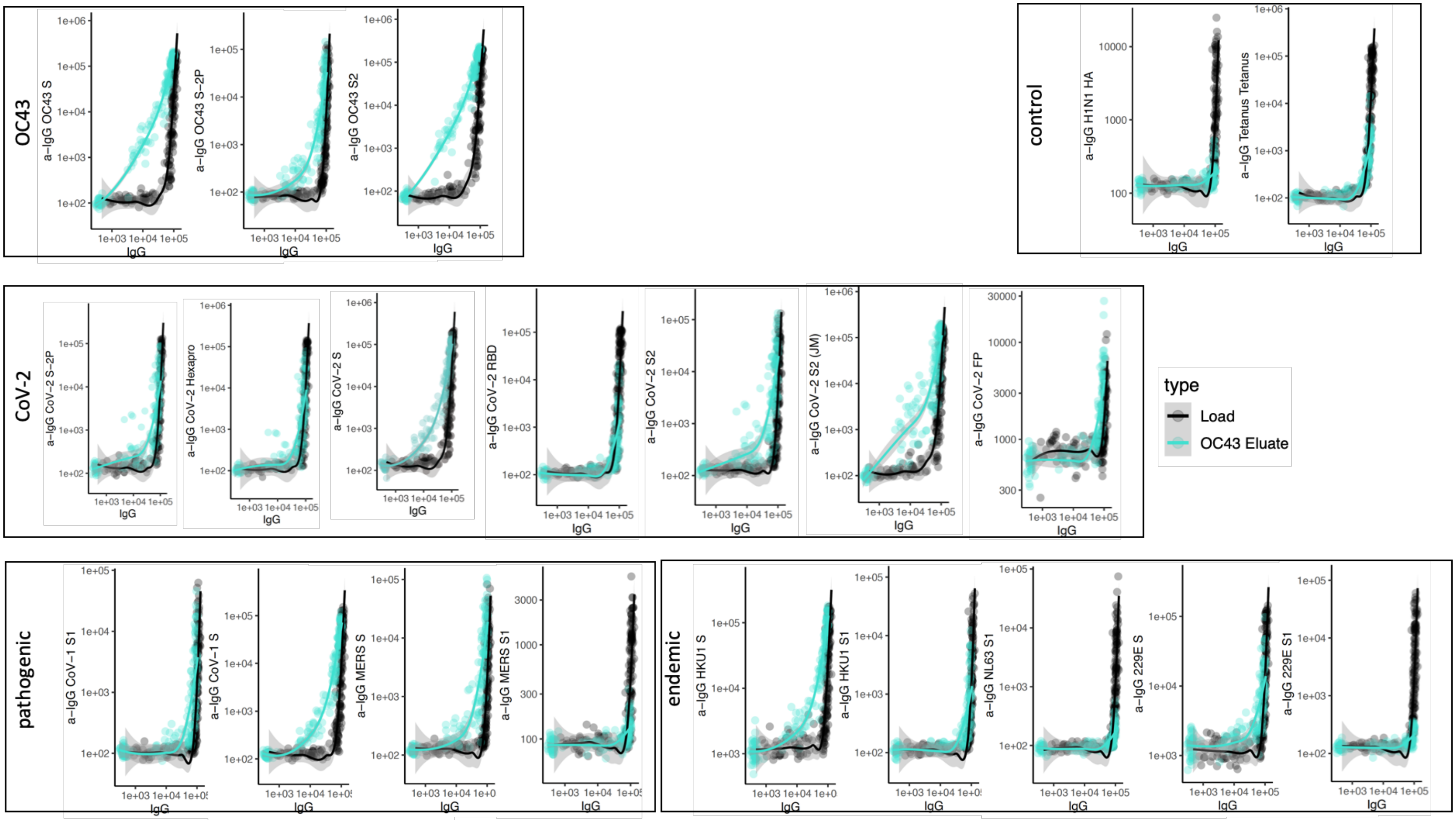


**Supplemental Figure 10: Cross-reactivity of OC43 S specific IgG.** Antigen binding profiles of IgG in unfractionated serum (load, black) and affinity-purified OC43 S- (eluate teal) fractions from 30 SARS-CoV-2 convalescent subjects across OC43, control, CoV-2, pathogenic CoV, and other endemic CoV. Y-axis depicts binding signal for indicated antigen specificity, x-axis depicts signal from total IgG quantitation.

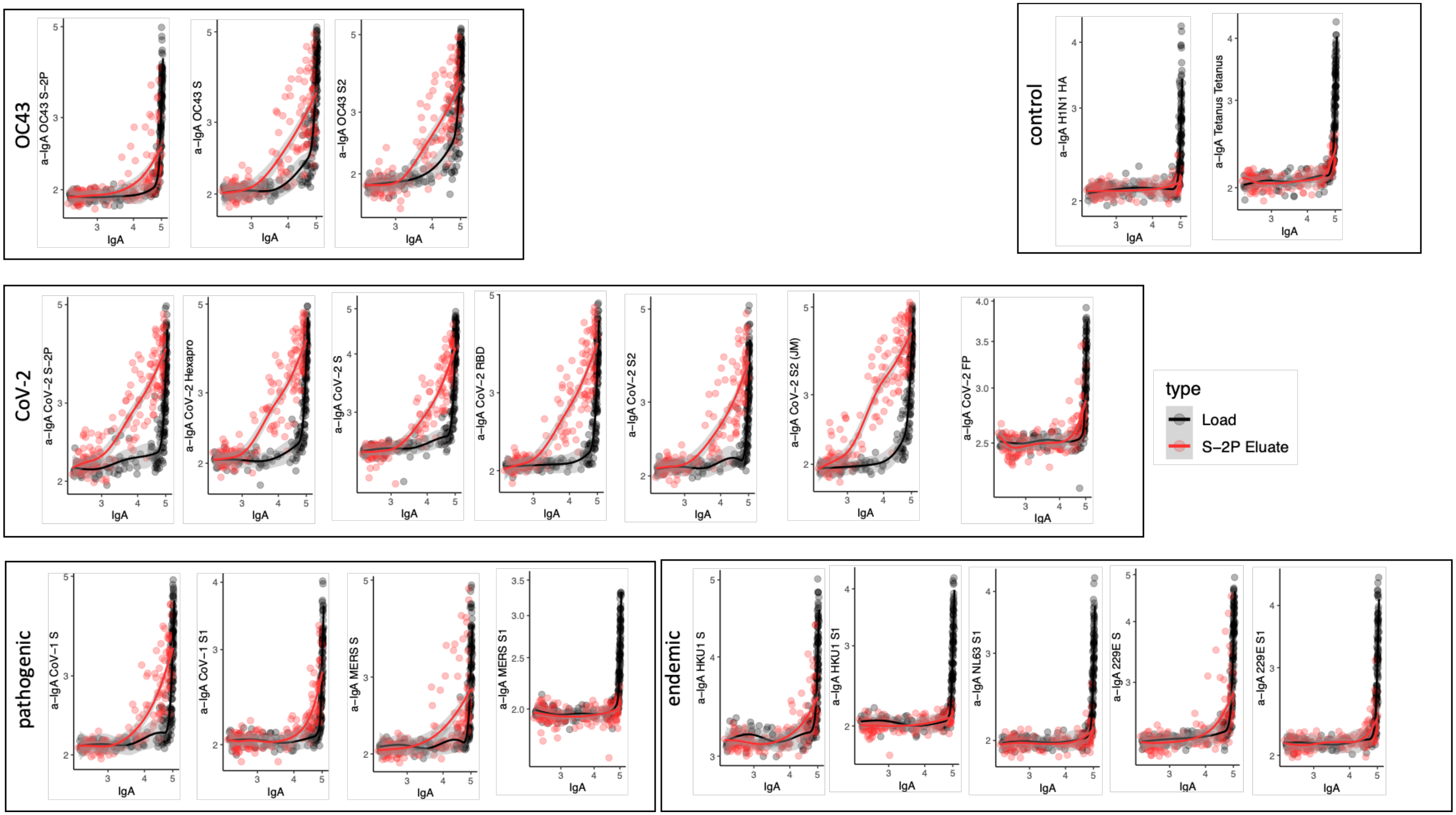


**Supplemental Figure 11: Cross-reactivity of SARS-CoV-2 S-2P specific IgA.** Antigen binding profiles of IgA in unfractionated serum (load, black) and affinity-purified CoV-2 S-2P- (eluate, red) fractions from 30 SARS-CoV-2 convalescent subjects across OC43, control, CoV-2, pathogenic CoV, and other endemic CoV. Y-axis depicts binding signal for indicated antigen specificity, x-axis depicts signal from total IgA quantitation.

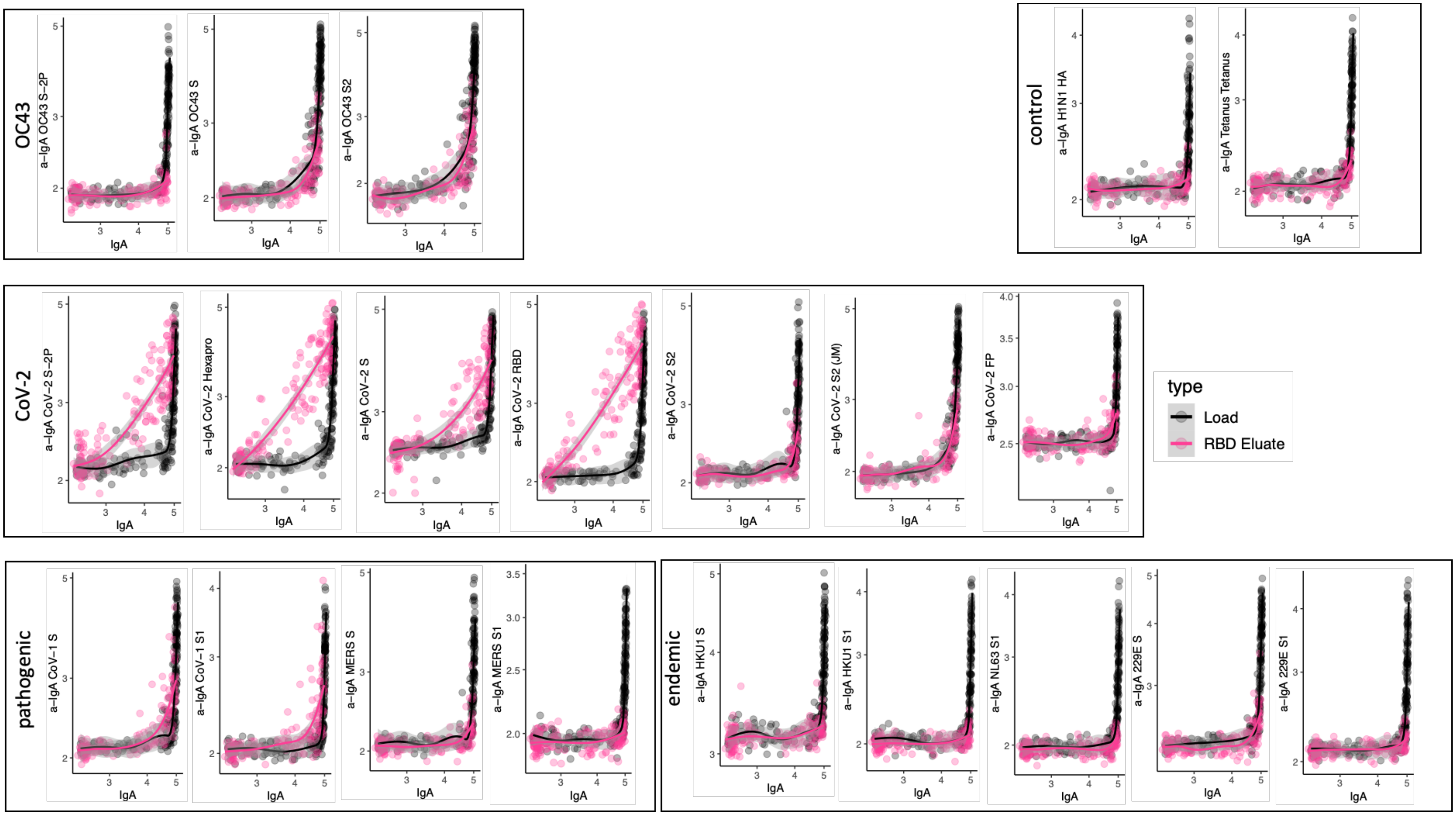


**Supplemental Figure 12: Cross-reactivity of SARS-CoV-2 RBD specific IgA.** Antigen binding profiles of IgA in unfractionated serum (load, black) and affinity-purified CoV-2 RBD- (eluate, pink) fractions from 30 SARS-CoV-2 convalescent subjects across OC43, control, CoV-2, pathogenic CoV, and other endemic CoV. Y-axis depicts binding signal for indicated antigen specificity, x-axis depicts signal from total IgA quantitation.

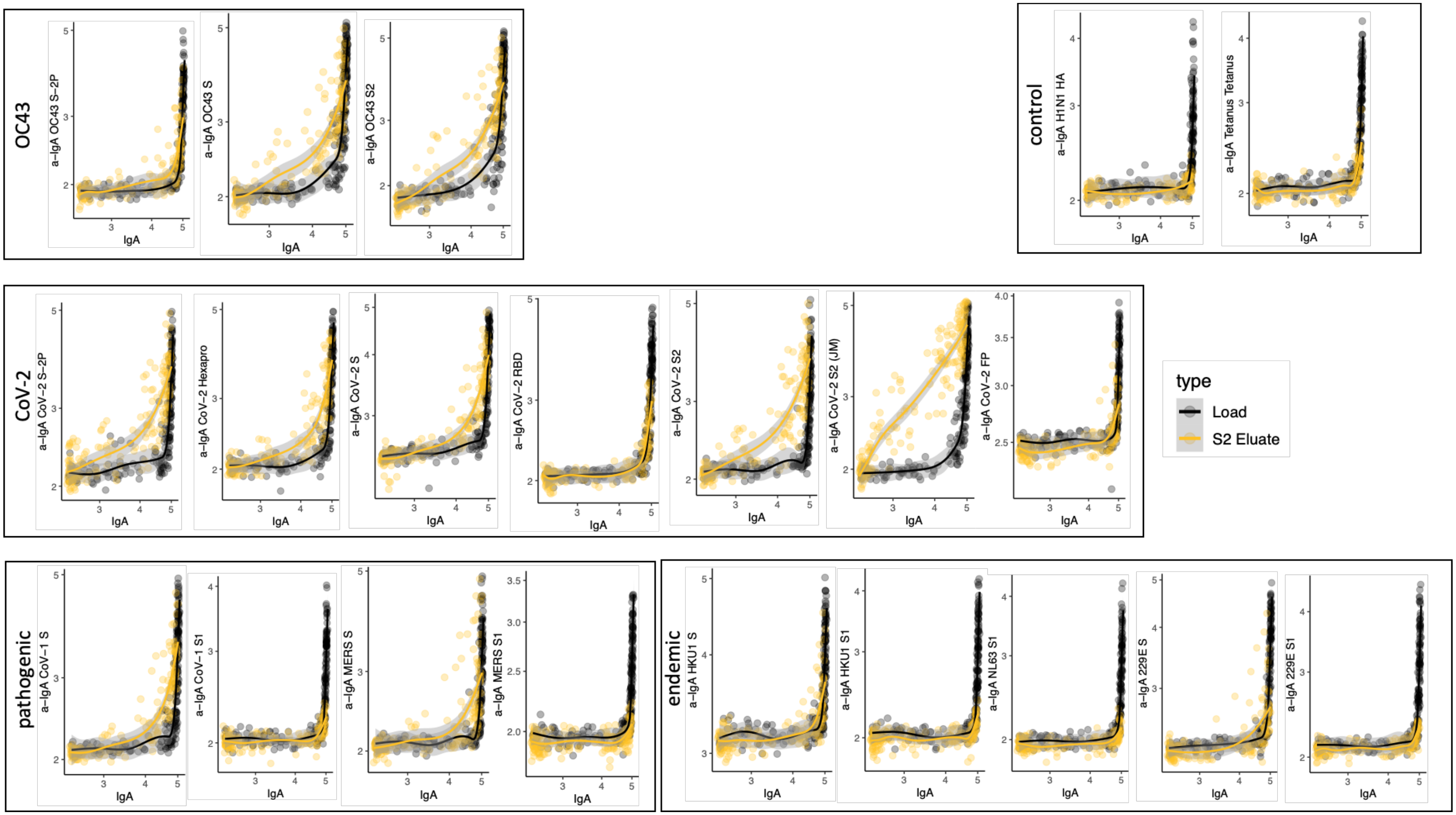


**Supplemental Figure 13: Cross-reactivity of SARS-CoV-2 S2 specific IgA.** Antigen binding profiles of IgA in unfractionated serum (load, black) and affinity-purified CoV-2 S2- (eluate, yellow) fractions from 30 SARS-CoV-2 convalescent subjects across OC43, control, CoV-2, pathogenic CoV, and other endemic CoV. Y-axis depicts binding signal for indicated antigen specificity, x-axis depicts signal from total IgA quantitation.

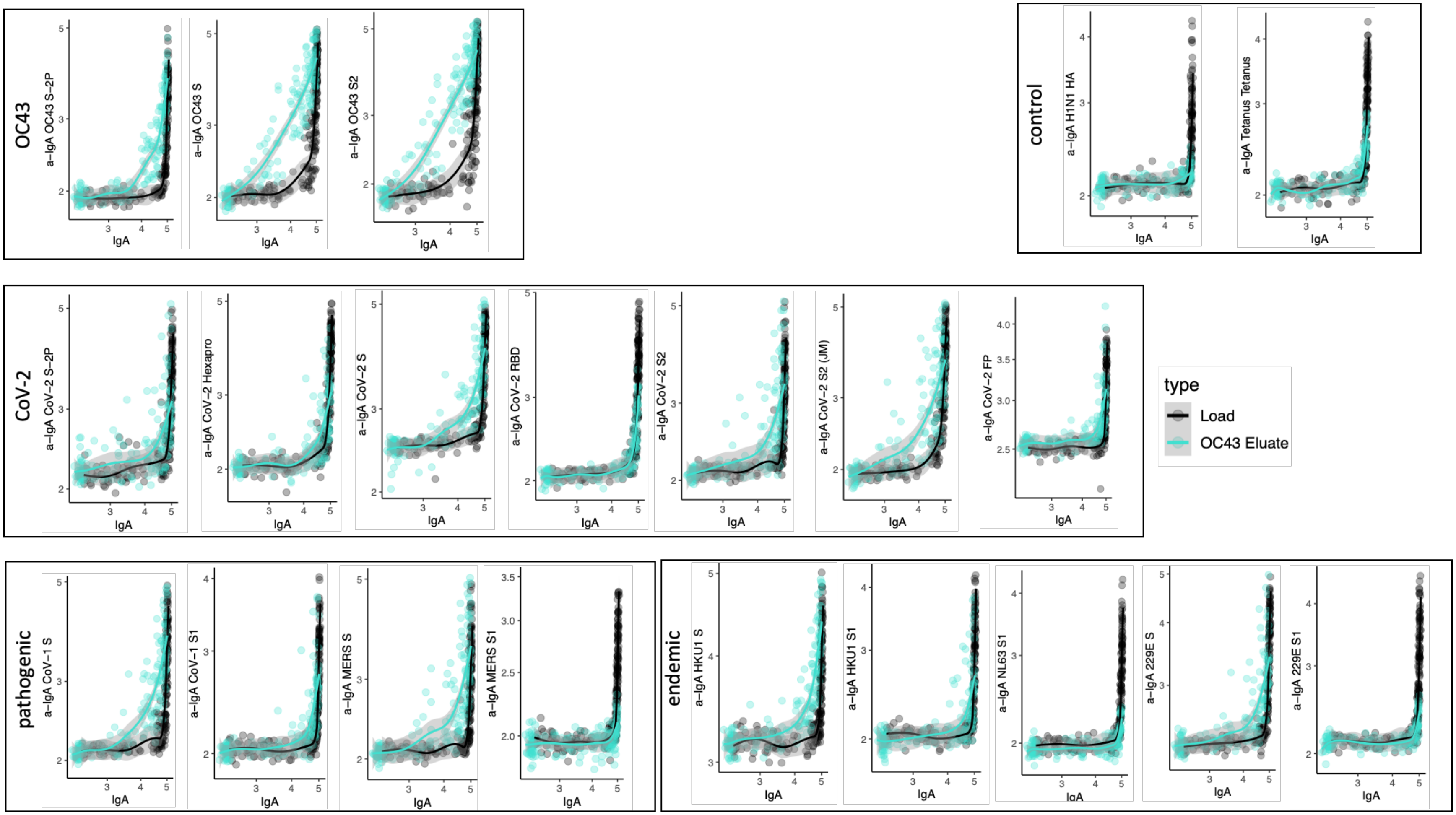


**Supplemental Figure 14: Cross-reactivity of OC43 S specific IgA.** Antigen binding profiles of IgA in unfractionated serum (load, black) and affinity-purified OC43 S- (eluate teal) fractions from 30 SARS-CoV-2 convalescent subjects across OC43, control, CoV-2, pathogenic CoV, and other endemic CoV. Y-axis depicts binding signal for indicated antigen specificity, x-axis depicts signal from total IgA quantitation.

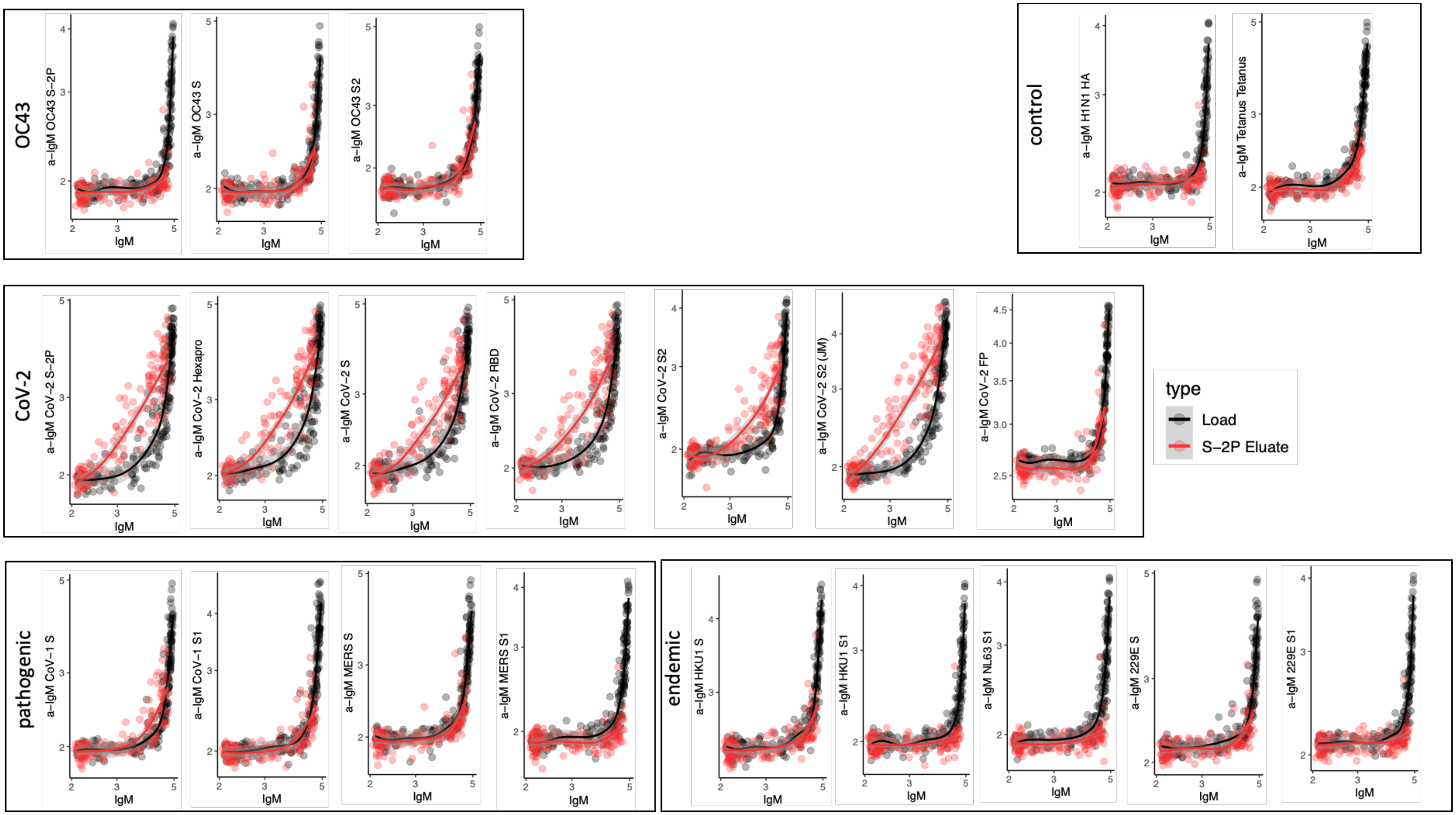


**Supplemental Figure 15: Cross-reactivity of SARS-CoV-2 S-2P specific IgM.** Antigen binding profiles of IgM in unfractionated serum (load, black) and affinity-purified CoV-2 S-2P- (eluate, red) fractions from 30 SARS-CoV-2 convalescent subjects across OC43, control, CoV-2, pathogenic CoV, and other endemic CoV. Y-axis depicts binding signal for indicated antigen specificity, x-axis depicts signal from total IgM quantitation.

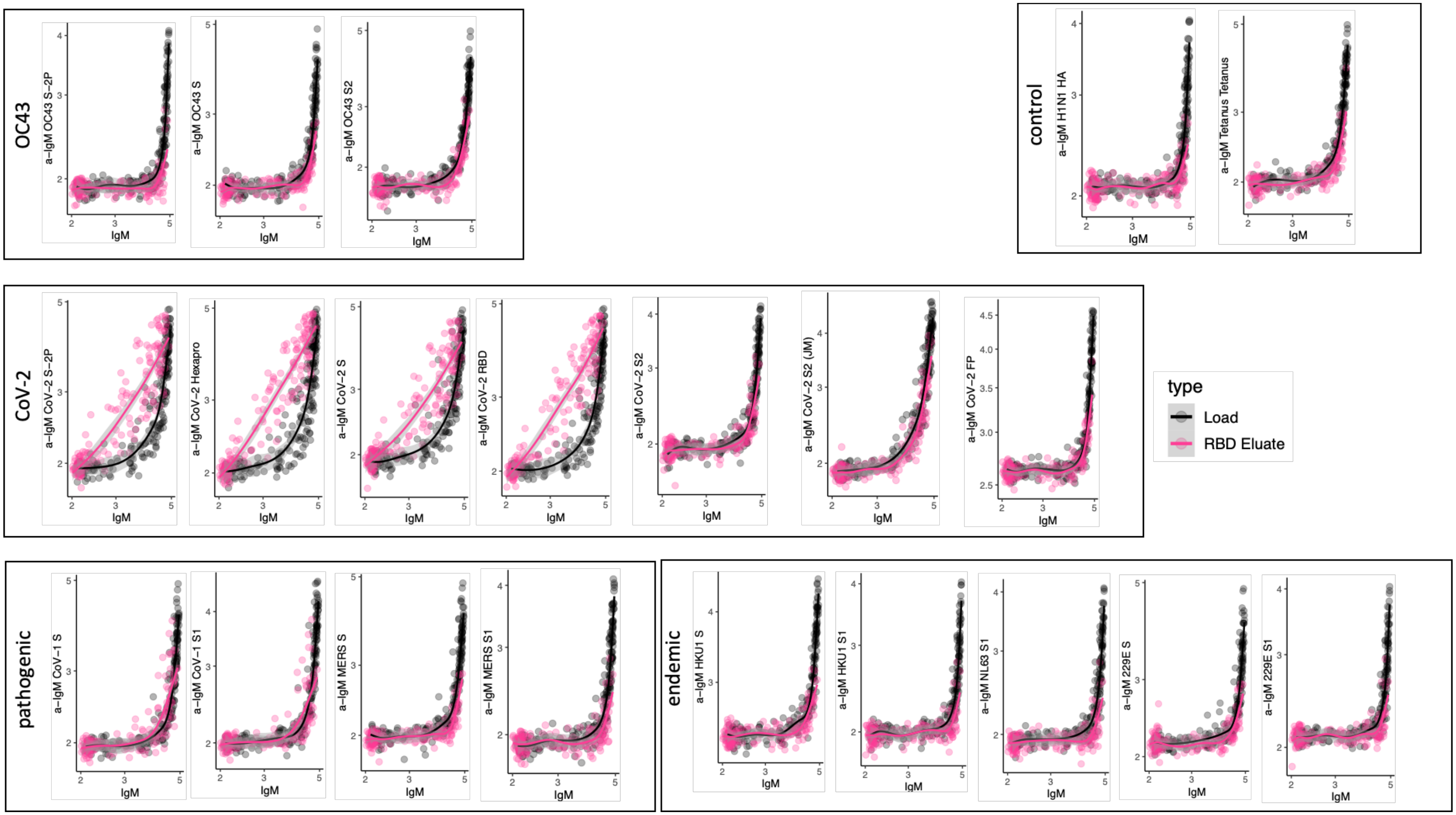


**Supplemental Figure 16: Cross-reactivity of SARS-CoV-2 RBD specific IgM.** Antigen binding profiles of IgM in unfractionated serum (load, black) and affinity-purified CoV-2 RBD- (eluate, pink) fractions from 30 SARS-CoV-2 convalescent subjects across OC43, control, CoV-2, pathogenic CoV, and other endemic CoV. Y-axis depicts binding signal for indicated antigen specificity, x-axis depicts signal from total IgM quantitation.

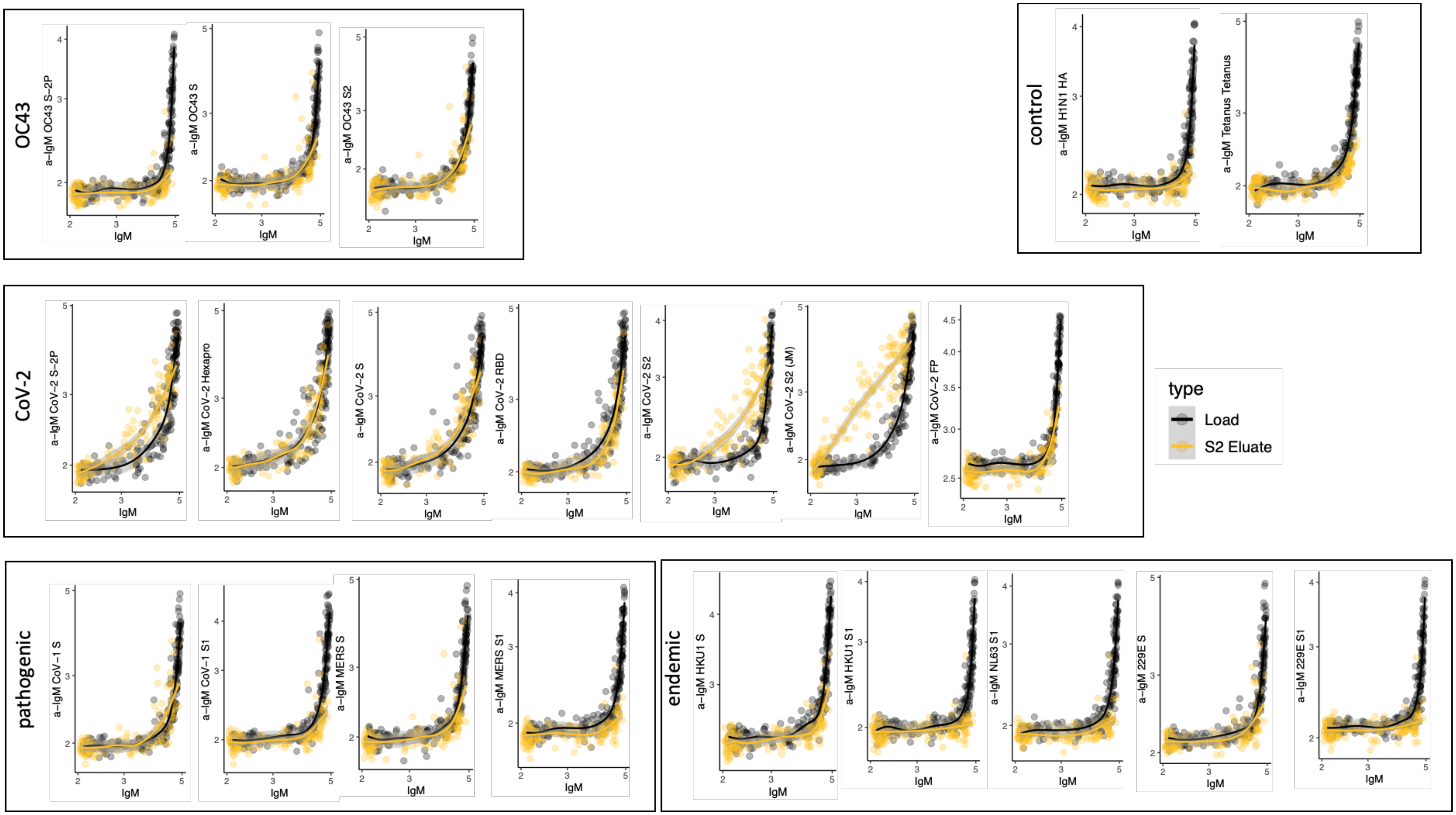


**Supplemental Figure 17: Cross-reactivity of SARS-CoV-2 S2 specific IgM.** Antigen binding profiles of IgM in unfractionated serum (load, black) and affinity-purified CoV-2 S2- (eluate, yellow) fractions from 30 SARS-CoV-2 convalescent subjects across OC43, control, CoV-2, pathogenic CoV, and other endemic CoV. Y-axis depicts binding signal for indicated antigen specificity, x-axis depicts signal from total IgM quantitation.

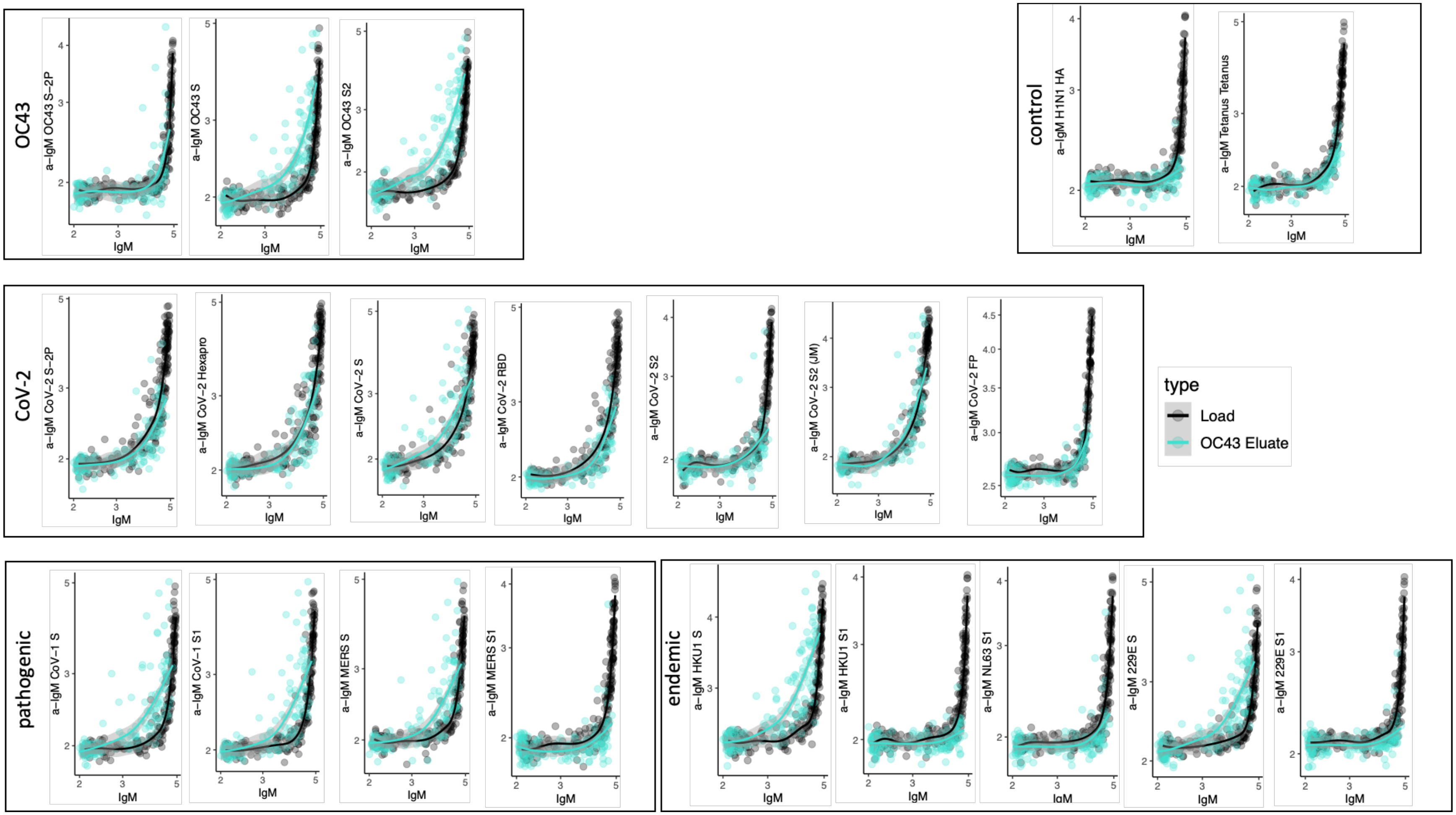


**Supplemental Figure 18: Cross-reactivity of OC43 S specific IgM.** Antigen binding profiles of IgM in unfractionated serum (load, black) and affinity-purified OC43 S- (eluate teal) fractions from 30 SARS-CoV-2 convalescent subjects across OC43, control, CoV-2, pathogenic CoV, and other endemic CoV. Y-axis depicts binding signal for indicated antigen specificity, x-axis depicts signal from total IgM quantitation.

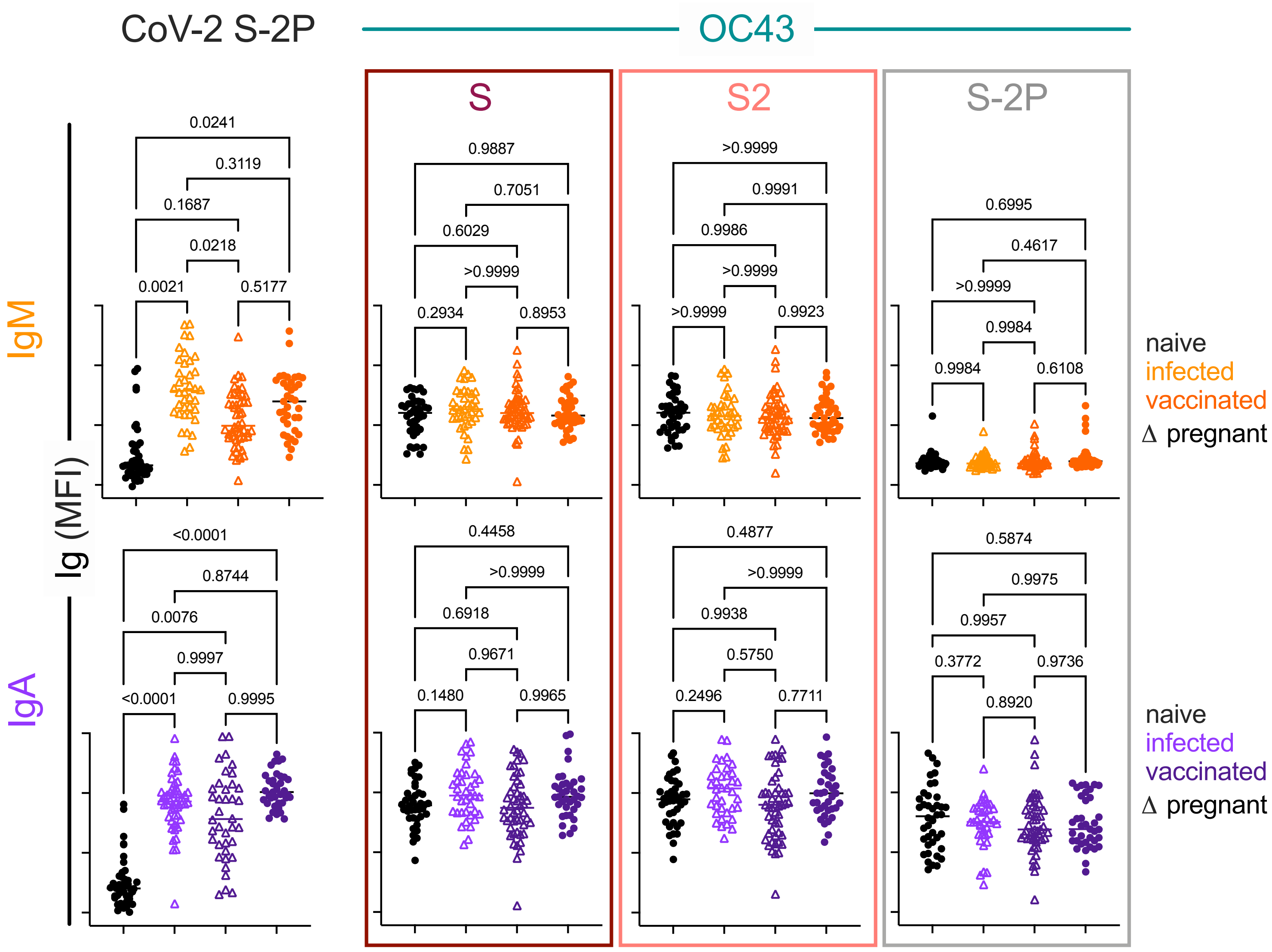


**Supplemental Figure 19. IgA and IgM responses among naïve subjects, infected pregnant women, vaccinated adults, and vaccinated pregnant women**. Box plots of IgM (top) and IgA (bottom) responses to SARS CoV-2 S-2P (left) and OC43 (right) S, S2, and S-2P. Statistical significance by ANOVA with Dunnett's correction.

**Supplemental Table 1**. Coronavirus structures. Spike protein information used to construct structural visualizations. PDB: Protein Data Bank; EM: electron microscopy.

| | PDB | Type | Resolution (Å) | Format |
|---|---|---|---|---|
| SARS-CoV-2 | 6XKL | EM | 3.21 | Trimer |
| 229E | 6U7H | EM | 3.1 | Trimer |
| OC43 | 6OHW | EM | 2.9 | Trimer |
| NL63 | 5SZS | EM | 3.4 | Trimer |
| HKU1 | 5I08 | EM | 4.04 | Trimer |
| SARS-CoV-2 Closed | 6X6P | EM | 3.22 | Trimer |

**Supplemental Table 2**. Fc detection and antigen reagents

| Antigen | Source |
|---|---|
| H1N1 HA1 | Immune Technology IT-003-00110p |
| HSV gE | Immune Technology IT-005-005p |
| Tetanus | Sigma 676570-37-9 |
| SARS CoV-2 N | Immune Technology IT-002-033Ep |
| SARS CoV-2 FP | New England Peptide |
| SARS CoV-2 S1 | ACRO Biosystems S1N-C52H3-100ug |
| SARS CoV-2 RBD | BEI Resources NR-52366 |
| SARS CoV-2 S2 | Immune Technology IT-002-034p |
| SARS CoV-2 S-2P | Expressed in Expi 293 |
| SARS-CoV-2 S-6P | Expressed in Expi 293 |
| WIV1 S-2P | Expressed in Expi 293 |
| SARS-CoV-1 S | Sino Biological 40634-V08B |
| SARS CoV-1 S1 | Sino Biological 40150-V08B1 |
| MERS S | Sino Biological |
| MERS S1 | Sino Biological 40069-V08B1 |
| OC43 S | Sino Biological 40607-V08B |
| OC43 S-2P | Expressed in HEK 293F |
| OC43 S2 | Sino Biological 40069-V08B |
| 229E S1 | Sino Biological 40605-V08H |
| 229E S | Sino Biological 40601-V08H |
| HKU1 S | Sino Biological 40606-V08H |
| HKU1 S1 | Sino Biological 40606-V08H |
| NL63 S1 | Sino Biological 40604-V08H |
| NL63 S | Sino Biological 40606-V08B |
| **Fc Detection** | **Source** |
| a- IgG | Southern Biotech 1030-09 |
| a-IgG1 | Southern Biotech 9054-09 |
| a-IgG2 | Southern Biotech 9070-09 |
| a-IgG3 | Southern Biotech 9210-09 |
| a-IgG4 | Southern Biotech 9200-09 |
| a-IgA | Southern Biotech 2050-09 |
| a-IgA1 | Southern Biotech 9130-09 |
| a-IgA2 | Southern Biotech 9140-09 |
| a-IgM | Southern Biotech 9020-09 |
